# Short-term effect of repeat echocardiographic screening for detection or confirmation of hypertrophic cardiomyopathy

**DOI:** 10.1101/2024.10.03.24314866

**Authors:** Mohammad Reza Movahed, Kyvan Irannejad, Fardokht Sadin, Luke Keating, Sharon S. Bates

## Abstract

**Background:** Diagnosis of Hypertrophic Cardiomyopathy (HCM) can be challenging. HCM can present later in life and the value of repeat echocardiogram for late presentation of HCM is not known. The goal of this study was to evaluate any changes in wall thickness occurring within 2 years using repeat screening echocardiography.

**Method:** The Anthony Bates Foundation has been performing screening echocardiography in high schools across the United States for the prevention of sudden death since 2001. A total of 206 subjects underwent baseline and repeat echocardiography within 2 years. We evaluated the prevalence of HCM before and after repeated echocardiography. Suspected HCM was defined as any wall thickness > 15 mm.

**Results:** The total prevalence of suspected HCM, defined by a cut-off value of 15 mm or more, was two from 206 (0.9%). Repeat echo within one year found one additional case of suspected HCM (wall thickness changed from 14 to 15 mm). However, the other two initial cases of suspected HCM showed regression of wall thickness to <15 mm not qualifying as HCM anymore as previously suspected after 2 years (one decreased wall thickness from 15 to 12 and the other one from 15 to 13 mm)

**Conclusion:** We found that repeat echocardiography is warranted in subjects with suspected HCM in order to prevent overdiagnosis. Furthermore, a new diagnosis of suspected cardiomyopathy was detected within 2 years of repeat screening. Larger studies are needed to evaluate the best timing and frequency of screening echocardiography for the detection and prevention of HCM.

## Introduction

Hypertrophic cardiomyopathy (HCM) is the most common inherited cardiovascular disease (1,2) and is the leading cause of sudden cardiac death (SCD) in young people, especially highly trained athletes (3–5). HCM is a common genetic cardiovascular occurring in every 500 individuals in the general population (6,7). Risk prediction models have estimated rates of SCD occurrences in childhood HCM within five years of diagnosis vary from 8% to 10% (8,9). HCM is defined by insufficient relaxation, hypercontractility, decreased compliance, and hypertrophy of the left ventricle (10–12). HCM is a chronic condition with a progressive course that can profoundly impact quality of life. Information on the societal cost linked to HCM has revealed substantial rises in hospitalizations for all causes, hospital stays, outpatient appointments, and overall healthcare expenses. The majority of cost increases are due to heightened hospitalizations and hospital days among symptomatic patients (13-15). Timely identification and accurate prognosis classification reduce the morbidity and mortality linked to this disease by facilitating prompt intervention, which can prevent SCD and other negative outcomes (16). Despite advancements, HCM remains underdiagnosed. While the population prevalence of HCM ranges from 1 in 200 to 1 in 500, only 10–20% of cases are identified through clinical methods. Although clinical and electrocardiographic information plays an important role, echocardiography remains the cornerstone in assessing patients with HCM. (17)

The diagnosis of HCM relies on imaging techniques such as echocardiography or cardiovascular magnetic resonance (CMR), which demonstrate an increase in the thickness of the left ventricular (LV) wall. HCM diagnosis has traditionally depended on the detection of left ventricular hypertrophy (LVH) with a wall thickness exceeding 15 mm using echocardiography or CMR, in the absence of any other cardiac or systemic disease (18). Patients with HCM need regular follow-up echocardiograms as per the 2020 AHA/ACC Guideline for the Diagnosis and Treatment of HCM (12). These echocardiograms are recommended every 1 to 2 years for asymptomatic patients to monitor any changes in the condition. Furthermore, a specific echocardiographic protocol for HCM should be used during follow-up appointments or if there is a change in the patient’s clinical status (19) (10). Follow-up echocardiography is crucial in the management of HCM. It is usually performed to evaluate the development of the condition and its effect on the heart as time progresses (20) (21)

Echocardiography allows for ongoing assessment of the heart’s structure, including the thickness of the heart muscle, left ventricular size, and heart chamber function. Follow-up echocardiography aids in evaluating and monitoring the presence and severity of left ventricular outflow tract obstruction, a common complication in HCM. The results of follow-up echocardiography can inform treatment decisions, such as the need for medications, lifestyle adjustments, or surgical interventions like septal myectomy. The timing and frequency of follow-up echocardiography will vary based on individual factors like symptom severity, previous imaging findings, and disease progression, typically recommended at regular intervals to monitor progress and treatment response. (10,20,21) However, data on the accuracy and reproducibility of repeat echocardiography in HCM screening are limited. Therefore, the aim of this study was to assess alterations in left ventricular wall thickness over a period of 2 years through repeat echocardiography in a young, asymptomatic sample presenting for screening.

### Method

The Anthony Bates Foundation has been performing screening echocardiography in high schools across the United States for the prevention of sudden cardiac death since 2001. A total of 206 subjects underwent baseline and repeat echocardiography within 2 years. We evaluated the prevalence of HCM before and after repeated echocardiography. Suspected HCM was defined as any left ventricular wall thickness > 15 mm.

### Results

The total prevalence of suspected HCM, defined by a cut off value of 15 mm or more, were two from 206 (0.9%). Rates of HCM identified on initial and repeat echocardiography are presented in Table 1. Repeat echo within one year found one additional case of suspected HCM (wall thickness changed from 14 to 15 mm). However, the other two initial cases of suspected HCM showed regression of wall thickness to < 15 mm not qualifying as HCM anymore as previously suspected after 2 years (one patient had decreased wall thickness from 15 mm to 12 mm and the other from 15 mm to 13 mm). Changes from initial to repeat echocardiography among the three patients who met guidelines for suspected HCM at any point are presented in Figure 1.

**Table 1.** Proportions of Positive HCM Screening and Repeat Echocardiography.

|  |  |  |
| --- | --- | --- |
|  | Screening | Repeat |
| Positive Screenings | $n = 2$ (0.9%) | $n = 1$ (0.005%) |
| Total Screenings | $n = 206$ | $n = 206$ |

**Figure 1.**
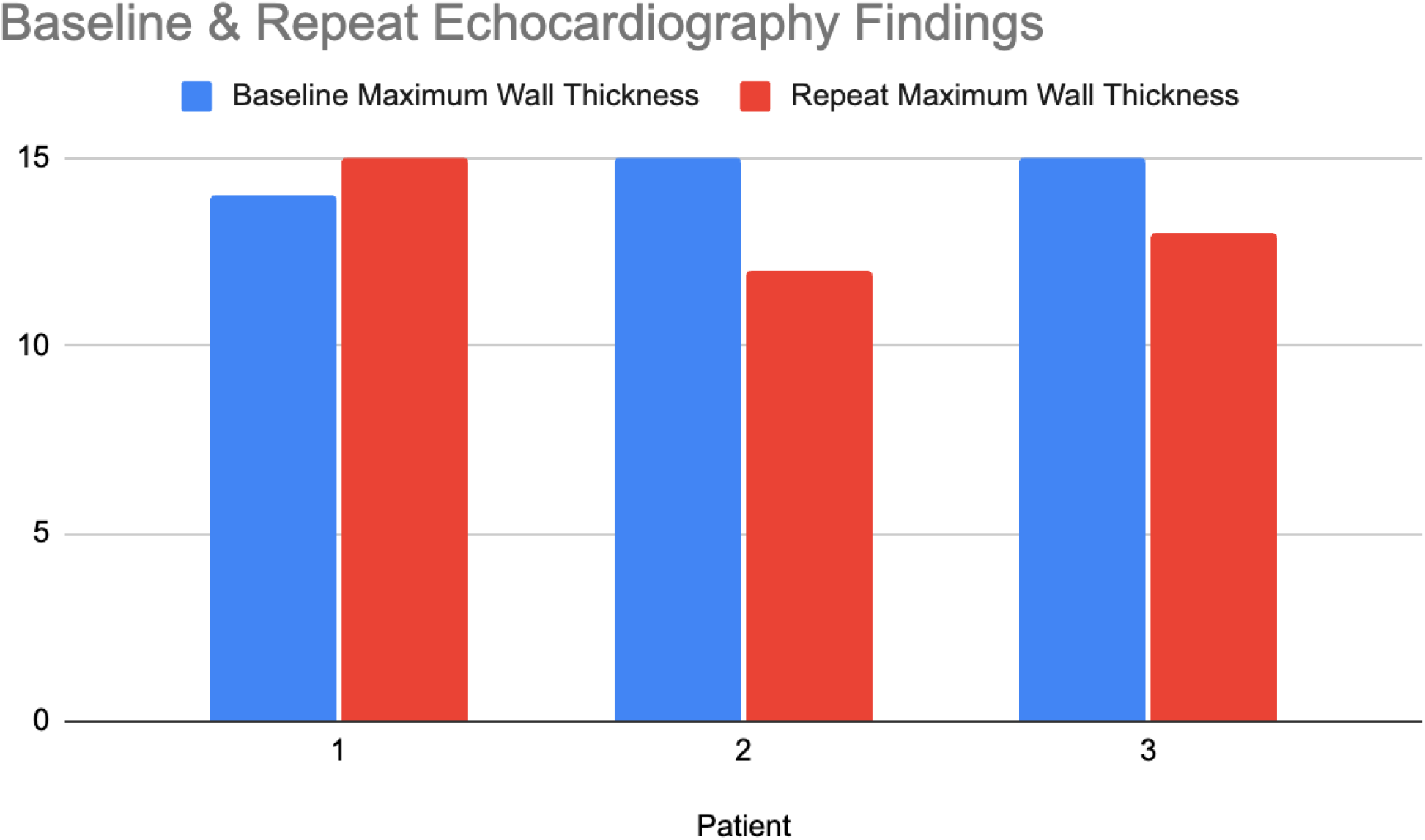

## Discussion

The current study found a low rate of identified HCM on echocardiography on both initial screening and two-year follow-up repeat echocardiography, with three total positive cases across screening timepoints. Of these three patients, the two patients who met criteria for HCM on initial screening no longer met criteria on repeat echocardiography. In addition, one patient whose maximum wall thickness was initially within normal limits then exceeded the 15mm cutoff on repeat screening. These data hold important preliminary implications for the utility of repeat screening with echocardiography in individuals at risk for HCM and highlight potential for pitfalls including inaccurate classification of patients due to limitations in test-retest reliability.

Recent evaluations of repeat screening echocardiography have highlighted issues in the reliability of results. For example, one large multi-center study evaluating variability in maximum wall thickness assessments found high rates of variability among readers in MWT on echocardiography, and low rates of agreement with CMR (24). Variability in left ventricular wall thickness on echocardiography in this sample would have led to risk reclassification in almost 20%, highlighting the potential for clinical misclassification resulting from reproducibility issues. (22)

Data from our young, relatively small, general population screening sample are consistent with potential for misclassification of risk due to variability in wall thickness assessments and suggest potential utility of repeat assessment to ensure accurate diagnosis. To maximize the benefit of repeat assessment, further research is needed to understand contributors to variability over time in left ventricular wall thickness on echocardiography. Change on repeated evaluation may be accounted for by multiple sources of variability or may reflect true change. Data on the reliability of repeat assessment will be crucial to improving diagnostic accuracy. However, a recent review paper concluded that assessments of the accuracy and reproducibility of echocardiography parameters are infrequent (23).

Emerging research has documented promising results for new echocardiography technology in improving the accuracy of HCM identification. For example, a recent review paper has highlighted potential for three-dimensional echocardiography and strain imaging echocardiography to improve upon current methods in precisely identifying HCM (24). Further, machine learning based approaches developed to improve upon accuracy and consistency in diagnosis and risk stratification have shown promising results. For example, one study comparing machine learning based wall thickness measurement to human readers found evidence for superior test-retest reliability and diagnostic precision of the new machine learning based approach (25). Similarly, fully automated echocardiography has shown preliminary evidence of superiority to manual assessments across a range of clinical parameters (26).

Interpretation of these findings should be considered in the context of several limitations. Firstly, our sample was relatively small and had a low positive screening for HCM, limiting our ability to evaluate inter-rater reliability and/or test-retest reliability. Furthermore, echocardiography was performed in a selected population of volunteers that may not be representative of a general population. For example, individuals volunteering for screening may be more likely to be at higher risk due to involvement in high-level athletic competition or family history of HCM.

## Conclusion

Based on this small study, repeat early echocardiography is warranted in subjects with suspected HCM in order to prevent over diagnosis. Our data suggest that increases in left ventricular wall thickness can occur in subjects with borderline findings over a relatively short follow up window of 2 years. Larger studies are needed to evaluate the best timing and frequency of screening echocardiography for detection and prevention of HCM.

## Data Availability

Not available

